# Self-reported preferences for seasonal Daylight Saving Time meet fundamentals of human physiology: correlations in the 2018 Public Consultation by the European Commission

**DOI:** 10.1101/2024.03.01.24303549

**Authors:** José María Martín-Olalla, Jorge Mira

**Affiliations:** Universidad de Sevilla, Facultad de Física, Departamento de Física de la Materia Condensada, ES41012 Sevilla, Spain; Universidade de Santiago de Compostela, Facultade de Física, Departamento de Física Aplicada and iMATUS, ES15782 Santiago de Compostela, Spain

**Keywords:** observational studies, statistical modelling, DST, summer time, latitude, sleep deprivation, DST, circadian misalignment, chronotype

## Abstract

We thoroughly analyze the results to question 2 (individual preferences for cancelling or keeping the current clock regulations) from the 2018 Public Consultation on summertime arrangements (DST) conducted by the European Commission.

We reveal correlations in the shares of population for cancelling the regulations and the winter sunrise time (SRW) [*R*^2^ = 0.177; *p* = 0.03; *N* = 25] and the onset of human activity [*R*^2^ = 0.677; *p* = 5 × 10^*−*5^; *N* = 17].

The results are in line with the rationale behind the regulations in the range of latitude 35^*°*^ to 63^*°*^: larger values of SRW (larger latitude) brought larger shares against the regulations; and earlier onset of human activity relative to SRW brought larger shares against the regulations.

The shares for cancelling the regulation did not show correlations with time offset (position in time zone), thus challenging the current view within the circadian community.

---

The seasonal clock regulations (seasonal Daylight Saving Time DST) aim to optimize the start of the day(Martín-Olalla and Mira, 2024) by aligning social activities closer to sunrise.(Hudson, 1898) This promotes a greater use of outdoor recreation in the spring-summer evening. The practice is based on human physiology as sunrise (morning light) rather than solar noon is the primary stimulus for keeping us aligned with the 24 h day.

At intermediate latitudes, this creates a conflict between clocks and social schedules, which are tied to noon, and human physiology. The clock policy helps alleviate this conflict and, all else equal, is equivalent to a seasonal schedule: working hours from 9am to 5pm during the autumn-winter season when the morning light is delayed, and from 8am to 4pm (though still rendered as 9am to 5pm) during spring-summer, when the morning light arrives earlier.(Martín-Olalla and Mira, 2025) Many Western societies welcomed this schedule during the past century, soon after social schedules became ubiquitous.

The clock policy is currently challenged by some chronobiologists and physiologist due to slight societal and health effects associated with the circadian disruption at the transition dates,(Fritz *et al*., 2020; Janszky and Ljung, 2008; Martín-Olalla and Mira, 2023) and circadian misalignment during the period when clocks are set to DST, allegedly “out of sync” with the sun.(Johnson and Malow, 2022; Kantermann *et al*., 2007; Roenneberg *et al*., 2019) This is noted by the time offset: the difference between solar noon and midday.

Since the clock regulations impact every aspect of social life, their sustainability is determined by social preferences which translates into policies. Thus, the understading of the current preferences and the mechanisms that shape them are of the utmost importance. In this line we should not forget the long-standing sustainability of the current clock policy suggests that “the issue that caused inconvenience was the changing of the clocks rather than the application of summertime [and winter-time] per se.”(Parliament *et al*., 2017, p. 16) and that clock regulations may have helped to keep the alignment of working hours with the sunrise.(Martín-Olalla, 2022)

We present here a thorough analysis of question 2 from the 2018 Public Consultation on summertime arrangements conducted by the European Commission. This web-based, self-reported survey targeted the EU population, which at that time consisted of 28 Member States.(for Mobility and Transport, 2019) Question 2 asked the individual preference to keep or to cancel the current arrangement, providing feedback on the *statu quo* under which the respondent has been living in the recent past. We do not analyze the preference for permanent DST or for permanent ST (question 5) because replies to this question reflect preferences for hypothetical, unknown scenarios that may or may not sustain if implemented.

Public consultations are unique because respondents are invited, but not compelled, to respond. Thus, they reflect self-reported, quasi-spontaneous preferences. The net balance of choices is often biased towards the points of view of those who choose to respond. For example, in 2019 the province of Alberta (Canada) conducted a public consultation on this policy: a 91 % of the 141 000 respondents (3 % of the population) favored permanent DST instead of the current policy.(Antle *et al*., 2022) In 2021, a binding referendum on this policy was held and a narrow 50.2 % of the respondents (39 % of registered voters, 25 % of the population) favored the current policy. Understandably the non-binding consultation attracted those more sensitive to the topic, which were those more discomforted with the current practice. In contrast, the binding referendum, held during municipal elections, increased participation from people less concerned with the issue and, eventually, more supportive of the current policy.

Our study does not aim to predict the outcome of a potential referendum, which is impossible, but to reveal how the shares of people against and for the regulations were distributed across the Member States. Since the public consultation was a synchronous observational experiment, the stimuli that prompted people to report their quasi-spontaneous preference should have been similar and an analysis of the result may help identify them.

Morning light should have impacted these preferences. We take winter sunrise time (SRW), a social synchronizer(Martín-Olalla, 2018), as a proxy for morning light. It is also a proxy for the winter photoperiod (2 ×(*T*_0_*/*2 − SRW)), and the seasonal span of sunrise times from winter to summer (2 *×* (SRW *− T*_0_*/*4)). Here *T*_0_ = 24 h is Earth’s rotation period and SRW is measured relative to solar midnight and, thus, a function of latitude only.

We used geographical coordinates of populated places in Europe from GeoNames (https://www.geonames.org) to retrieve the SRW and the time offset (the difference between clock noon and solar noon) for each location. We then determined the population-weighted median SRW and time offset by Member State. We note that median SRW changes in Europe from 7.18 h (Cyprus) to 9.48 h (Finland) spanning 2.30 h, larger than the size of the clock changing. This variation makes the 2018 public consultation ideal to test the role of SRW in this topic. Table S1 in the supplementary material available at doi:10.5281/zenodo.14619659 lists geographical and societal values, see also figure S1.

We analyzed the results of the two choices presented in Question 2 (cancel or keep the current regulations) by Member State. We scaled the results using population numbers (obtained from Eurostat table doi:10.2908/DEMO·PJAN) and with the percentage of access to the internet (from the World Bank https://data.worldbank.org/indicator/IT.NET.USER.ZS). This allowed us to compute the shares of target population for canceling (*C*) and for keeping (*K*) the regulations. Save for Malta, the number of respondents is greater than 1000, thus *C* and *K* can be treated as continuous, independent variables. See Table S2.

The null hypothesis of normally distributed shares (*C* and *K*) did not hold at the standard level of significance (*α* = 0.05) with *p* = 0.02 (*C*) and *p* = 0.01 (*K*). We used Tukey’s fences (with a size *k* = 1.5) to remove outliers and retain observations within the bracket [*Q*_1_ −*k* (*Q*_3_− *Q*_1_), *Q*_3_ + *k* (*Q*_3_ −*Q*_1_)], where *Q*_1_ and *Q*_3_ are the low and high quartiles of the distribution. After removing outliers, the null hypothesis of normally distributed shares was sustained (*p* = 0.60 and *p* = 0.48) with averages 0.393 % (*C*) and 0.074 % (*K*) and relative standard errors 69 % and 83 % respectively.

In bivariate analysis (Table S3), the null hypothesis “*C* does not depend on SRW” failed to sustain (*R*^2^ = 0.177; *p* = 0.036; *N* = 25): Member States with larger SRW (larger latitude) were significantly more likely to have larger shares of population against the current policy in the Public Consultation. In contrast, the null hypothesis “*K* does not depend on SRW” sustained (*R*^2^ = 0.030; *p* = 0.420; *N* = 24) at the standard level of significance. When using the time offset as a descriptor, the null hypothesis was sustained for *C* and *K* (figure S4). The null hypotheses of normally distributed residuals sustained for all analyses.

Figure 1 shows scatter plots of *C* (top) and *K* (bottom) versus SRW with the same vertical scale. The lines display the bivariate linear regressions. The low rate of responses for keeping the regulations may be linked to the lack of interest of this group, mimicking the results of Alberta. Therefore, the uncontrolled confounding factors dominated the distribution of shares and resulted in no statistically significant correlations. In contrast, the shares of the opposing group were high enough to yield an association between *C* and SRW that was able to partially overcome the uncontrolled factors.

**Figure 1.**
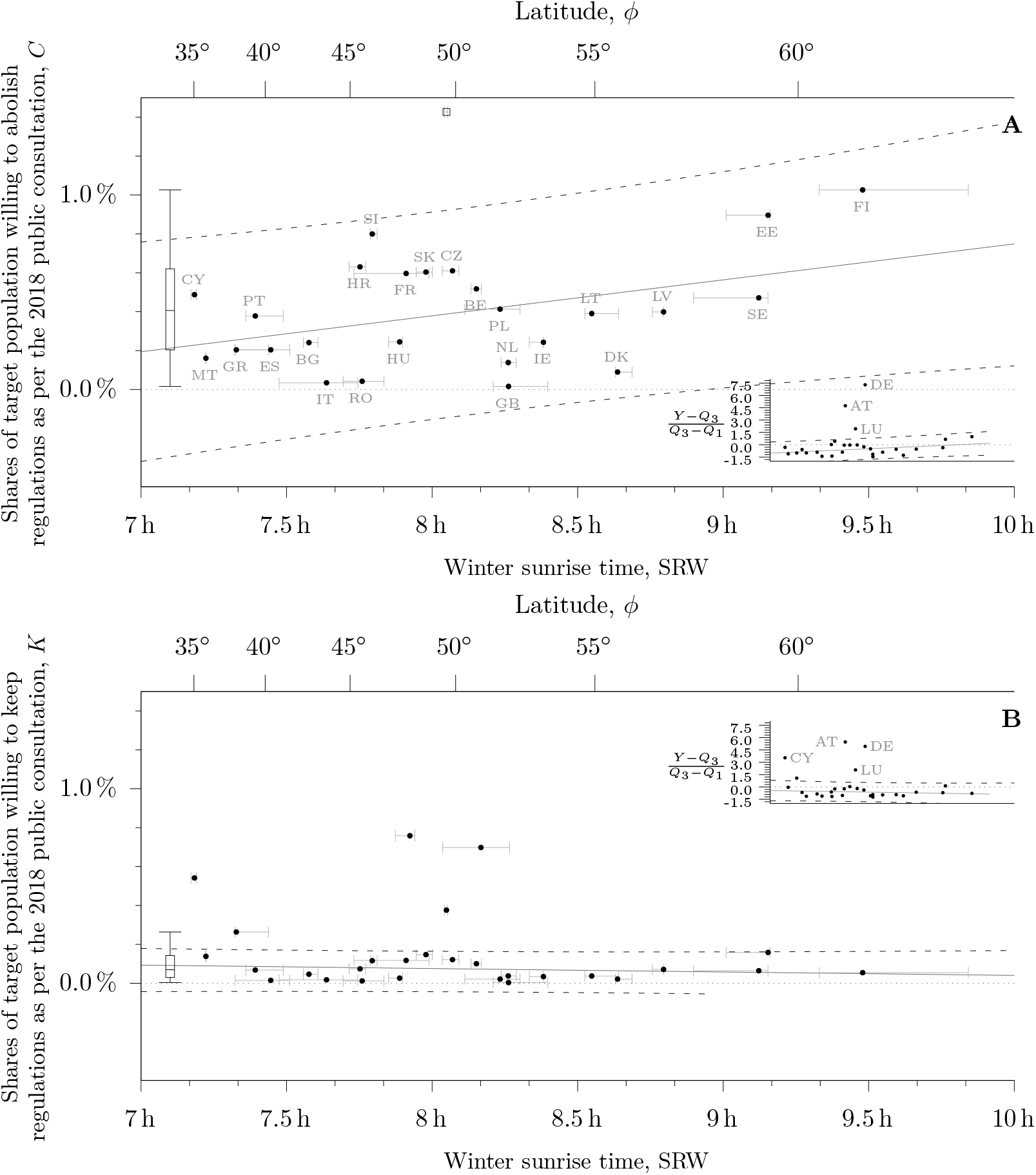
**A**. The association between the winter sunrise time SRW (solar time) and the shares of population willing to cancel DST as per the 2018 public consultation *C*. The inset shows the distribution of shares scaled by the interquartile range to identify and quantify the outliers Austria, Germany and Luxembourg, not shown in the main picture and excluded from the regression. The low quartile was *Q*_1_ = 0.204 % and the high quartile, *Q*_3_ = 0.620 % (see the boxplot on the left side of the plot). The solid line shows the point estimates of the prediction; the dashed lines bound the 95 % prediction interval. The regression is statistically significant (*p* = 0.03) at the standard level of significance (*α* = 0.05). Notice that the span of the predictor is larger than one hour: the standard unit of social time and the size of the shift brought by DST to clocks. Horizontal bars highlight population weighted low and high quartiles (France and Spain show the span of the European areas only). **B**. The same picture but for the shares of population willing to keep regulations as per the 2018 public consultation. The association does not bring statistically significant correlations. For the shares *K, Q*_1_ = 0.030 % and *Q*_3_ = 0.143 % (see the boxplot on the left side of the plot). Panels a A and B have the same vertical scale for the sake of comparison, a close up of panel B is available in figure S5. Labels (iso-3166 alpha-2) in increasing values of latitude: CY Cyprus; MT Malta; GR Greece; PT Portugal; ES Spain; BG Bulgaria; IT Italy; HR Croatia; RO Romania; SI Slovenia; HU Hungary; FR France; AT Austria; SK Slovakia; LU Luxembourg; CZ Czech Republic; BE Belgium; DE Germany; PL Poland; GB United Kingdom; NL Netherlands; IE Ireland; LT Lithuania; DK Denmark; LV Latvia; SE Sweden; EE Estonia; FI Finland.

For the reader’s interest we also analyzed combinations of *C* and *K*. The net balance *C* −*K* resulted in a statistically significant correlation (*R*^2^ = 0.247; *p* = 0.010; *N* = 26, see figure S6), with a larger net balance as SRW increases. The ratio *C/K* also yielded statistically significant correlation (*R*^2^ = 0.239; *p* = 0.013; *N* = 25, figure S7). We note that Alberta’s *C/K* agrees well with EU results. In contrast, the turnout *C* + *K* showed a statistically non-significant correlation (*R*^2^ = 0.057; *p* = 0.252; *N* = 25; figure S8) because the increase of *C* with SRW and the decrease of *K* with SRW offset each other. We took specific Tukey’s fences associated with each combination.

The clock policy aims to regulate the onset of social activity after the night period of rest. To provide more insight we draw attention to the Harmonized European Time Use Survey (HETUS)(Eurostat, 2010) which provides the daily rhythm of sleep and other personal cares, as well as the daily rhythm of work by Member State. We compute the onset time *t* as the moment in the morning when the daily rhythm overtakes (for work) or undertakes (for sleep) half of the maximum value of the daily rhythm. We then compute the distance *t*_*w*_ = *t* − SRW to the winter sunrise time and the distance to the solar noon *t*_*s*_ (table S4 and figure S3).

We correlated *t*_*w*_ with the shares *C* in the *N* = 17 Member States that have reported daily rhythm to Hetus and found that the null hypotheses “*C* does not depend on *t*_*w*_” did not sustain at the standard level of significance *R*^2^ = 0.542; *p* = 5 *×*10^*−*5^; *N* = 17; table S5 (for work) and *R*^2^ = 0.364; *p* = 0.008; *N* = 17; table S5 (for sleep and other personal cares). Member States with earlier onset of social activity, relative to SRW, were more likely to yield larger shares of population against the regulations in the Public Consultation. Figure 2 shows a scatter plot of *C* versus sleep offset (top) and work onset (bottom), and the bivariate linear regressions.

**Figure 2.**
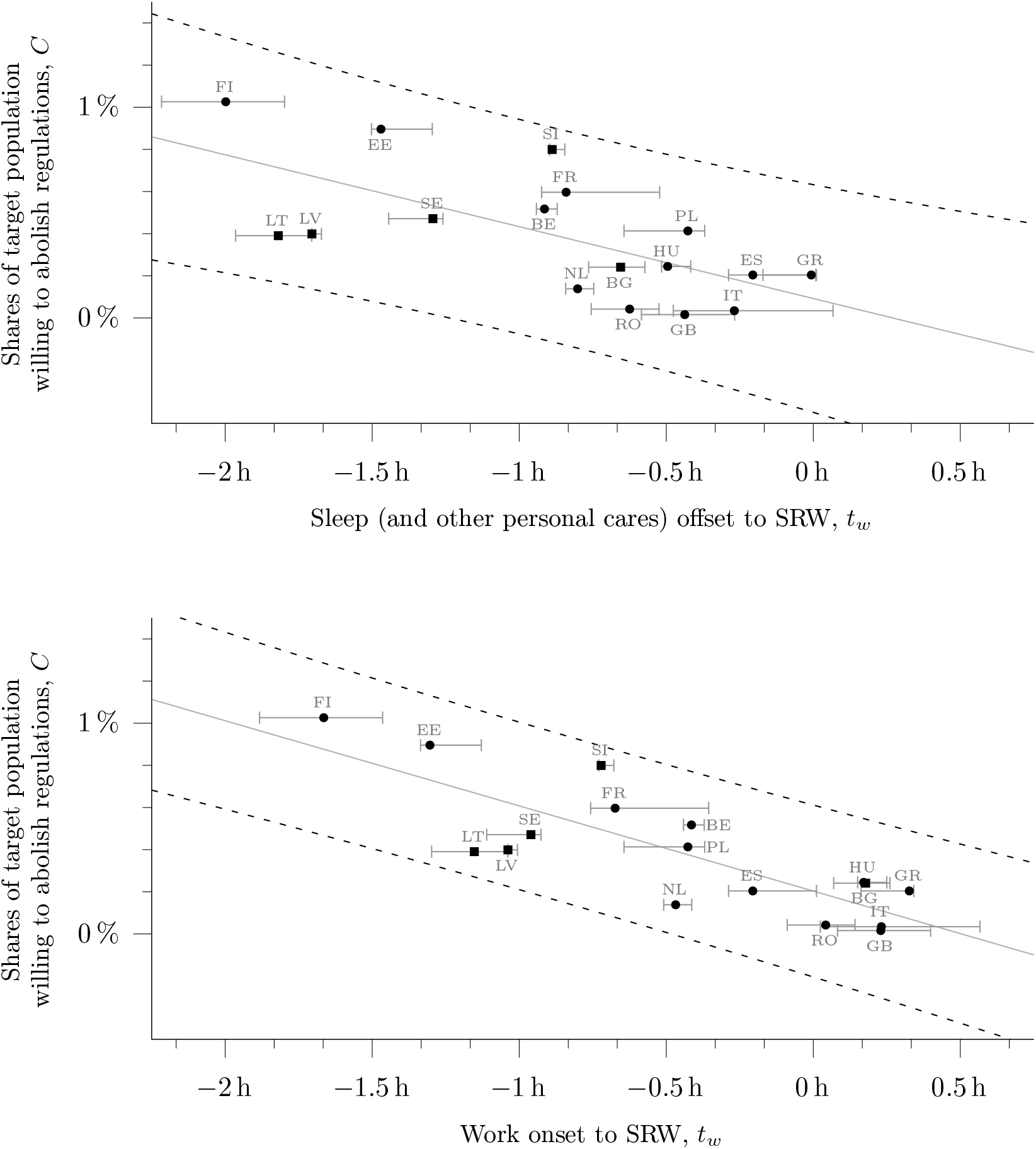
Scatter plots of the shares of target population willing to cancel DST as per the 2018 European Commission public consultation (outcome) vs two predictors related to human activity as per HETUS: sleep offset to SRW (top) and work onset to SRW (bottom). Circles refer to round 2 (year 2010) of HETUS, and squares to round 1 (year 2000). The null hypothesis “the predictor does not explain the outcome” does not sustain at the standard level of significance for either test and brought larger shares *C* with earlier onset/offset relative to SRW. Solid lines show the results of the linear regression; dashed lines bound the 95 % prediction interval of the regression. Horizontal bars highlight population weighted low and high quartiles (France and Spain show the span of the European areas only). The null hypotheses sustained if the descriptor is set to distance to noon.

There are many limitations in this study connected to the fact that the public consultation was an uncontrolled, observational experiment without sample design. As an example 70 % of the replies came from Germany, which accounts for only 15 % of the population. Germany, Austria and Luxembourg are outliers for *C* and *K* in this study and are the only Member States where German is the official language or is widely spoken by the general population. This suggests that German-language media may have given more attention to the public consultation than elsewhere. Since they are not outliers for *C/K*, the extra attention should have been unbiased.

Likewise, Poland and Finland were outliers in the ratio *C/K*: these Member States initiated the push for canceling the regulations, which might be reflected by this statistics. Spain was an outlier in this metric. We link this to the fact that some lobbyist associations argue that Spain is in the “wrong time zone”, a claim further exacerbated during DST,(Roenneberg *et al*., 2019) when clock time is 2 h ahead of mean solar time. This claim is a misunderstanding since time zones are arbitrary conventions that, by themselves, are neither wrong nor right. Life in Spain continues to be dictated by the natural cycle of light and dark, in sync with the sun,(Martín-Olalla, 2018) but some population groups are sensitive to these false beliefs.

The sample sizes, represented by the average shares of target population 0.394 % (*C*) and 0.074 % (*K*), are low to predict the net result of a would-be referendum, as exemplified by the case of Alberta. However, the median shares are high enough for observational studies. As an example Kantermann *et al*. (2007) was an observational, self-reported study whose sample size was *∼* 0.09 % of the German population.

It is important to note that confounding, uncontrolled factors seldom if ever build up correlations. The significance of *p* = 5 × 10^*−*5^ in the bivariate correlations between the work onset distance to SRW and *C* indicates that there are 5 in 100 000 odds that the correlation arose by chance. Thus, the trends in the 2018 public consultation highlight the role played by the morning light (SRW) in shaping quasi-spontaneous preferences for the current regulations and cannot be explained by the prevailing view within the circadian community, which associates clock regulations —and their discomfort— to time offset.

The results also reflect a simple physiological response to external stimuli. Early risers do not need a further advance of the activity in spring, they find dark morning conditions after DST onset and before DST offset, and their summer bedtimes come close to summer sunset. All these factors contribute to build up a discomfort for the practice. Conversely, late risers tend to report less discomfort for the current regulations because they welcome an advance of their late activity when the environmental conditions allow it —they welcomed this in the 1920s—, and are not challenged by light conditions after or before transition dates.

In shaping the preferences for clock regulations we do not exclude the interplay of other health or physiological issues associated, for instance, with individual chronotype. Our analysis just emphasizes the importance of environmental (latitude) and societal (onset times) conditions that underpin the clock regulations. Any choice —whether advancing schedules, delaying schedules, or the current alternating schedules— would bring discomfort to certain population groups. The intriguing question that arises is whether the current regulations are still the best practical compromise that modern societies regulated by schedules can achieve beyond 30^*°*^ latitude to minimize the discomfort for both early risers and late risers.

The Authors declares no competing interests.

## Supporting information

Supplemental tabular and graphical material

## Data Availability

All data produced in the present work are contained in the manuscript.
A data-set is accesible at
https://doi.org/10.5281/zenodo.13301201

## REFERENCES

Antle, Michael C, Mahtab Moshirpour, Patricia R Blakely, Katelyn Horsley, Colin J Charlton, and Victor Hu (2022), “Longitudinal location influences preference for daylight saving time.” Journal of biological rhythms 37, 343–348.

Eurostat, (2010), “Harmonised european time use surveys (hetus). rounds 1 and 2,”.

Fritz, Josef, Trang VoPham, Kenneth P. Wright, and Céline Vetter (2020), “A chronobiological evaluation of the acute effects of daylight saving time on traffic accident risk,” Current Biology 30, 729–735.

Hudson, George Vernon (1898), “On seasonal time,” Transactions and Proceedings of the New Zealand Royal Society 31, 577–598.

Janszky, Imre, and Rickard Ljung (2008), “Shifts to and from daylight saving time and incidence of myocardial infarction,” New England Journal of Medicine 359, 1966–1968.

Johnson, Karin G, and Beth A. Malow (2022), “Daylight saving time: Neurological and neuropsychological implications,” Current Sleep Medicine Reports 8, 86–96.

Kantermann, Thomas, Myriam Juda, Martha Merrow, and Till Roenneberg (2007), “The human circadian clock’s seasonal adjustment is disrupted by daylight saving time,” Current Biology 17, 1996–2000.

Martín-Olalla José María (2018), “Latitudinal trends in human primary activities: characterizing the winter day as a synchronizer,” Scientific Reports 8, 5350.

Martín-Olalla José María (2022), “A chronobiological evaluation of the risks of canceling daylight saving time,” Chronobiology International 39, 1–4.

Martín-Olalla José María, and Jorge Mira (2023), “Sample size bias in the empirical assessment of the acute risks associated with daylight saving time transitions,” Chronobiology International 40, 186–191.

Martín-Olalla José María, and Jorge Mira (2024), “Assessing the best hour to start the day: an appraisal of seasonal daylight saving time,” zenodo 10.5281/zenodo.13990240.

Martín-Olalla José María, and Jorge Mira (2025), “Seasonal daylight saving time in uk: a long-standing, successful record with few reasons to alter,” Journal of Sleep Research 34, e14420.

for Mobility, European Commission DG, and Transport (2019), Technical assistance with the public consultation on EU summertime arrangements: final report (Publications Office).

Parliament, European, Directorate-General for Parliamentary Research Services, and Irmgard Anglmayer (2017), EU summer-time arrangements under Directive 2000/84/EC Study (European Parliament).

Roenneberg, Till, Eva C. Winnebeck, and Elizabeth B. Klerman (2019), “Daylight saving time and artificial time zones – a battle between biological and social times,” Frontiers in Physiology 10, 944.

