## Supplemental tabular and graphical material for "Self-reported preferences for seasonal Daylight Saving Time meet fundamentals of human physiology: correlations in the 2018 Public Consultation by the European Commission"

This is a supplemental information file.
This version is published in medR$\chi$iv under doi:10.1101/2024.03.01.24303549

### Supplemental information to: Self-reported preferences for Daylight Saving Time meet fundamentals of human physiology: correlations in the 2018 Public Consultation by the European Commission

José María Martín-Olalla

*Universidad de Sevilla, Facultad de Física, Departamento de Física de la Materia Condensada, ES41012 Sevilla, Spain*[*]

Jorge Mira

*Universidade de Santiago de Compostela, Facultade de Física, Departamento de Física Aplicada and iMATUS, ES15782 Santiago de Compostela, Spain*[†]

(Dated: January 11, 2025)

Keywords: observational studies; quantile regression; statistical modelling; bivariate regression; DST; summer time; society; latitude; sleep deprivation; spring transition; Europe; America; season

Typeset by REVTEX

#### 1. TABULAR MATERIAL

| Country | | Latitude $\phi$ | Longitude $\lambda$ | Time offset $Z-\lambda/\Omega$ | Winter sunrise, SRW | Population $P$ | Internet usage $I$ |
|---|---|---|---|---|---|---|---|
| CY | Cyprus | 35.0° | 33.4° | −0.22 h | 7.18 h | 864 236 | 84.43 % |
| MT | Malta | 35.9° | 14.5° | 0.04 h | 7.22 h | 475 701 | 81.66 % |
| GR | Greece | 38.1° | 23.6° | 0.42 h | 7.33 h | 10 741 165 | 72.24 % |
| PT | Portugal | 39.3° | −8.7° | 0.58 h | 7.39 h | 10 291 027 | 74.66 % |
| ES | Spain | 40.4° | −3.6° | 1.20 h | 7.45 h | 46 658 447 | 86.11 % |
| BG | Bulgaria | 42.7° | 24.8° | 0.35 h | 7.58 h | 7 050 034 | 64.78 % |
| IT | Italy | 43.7° | 12.2° | 0.19 h | 7.64 h | 60 483 973 | 74.39 % |
| HR | Croatia | 45.5° | 16.1° | −0.07 h | 7.75 h | 4 105 493 | 75.29 % |
| RO | Romania | 45.6° | 25.6° | 0.29 h | 7.76 h | 19 533 481 | 70.68 % |
| SI | Slovenia | 46.2° | 14.6° | 0.03 h | 7.79 h | 2 066 880 | 79.75 % |
| HU | Hungary | 47.5° | 19.0° | −0.27 h | 7.89 h | 9 778 371 | 76.07 % |
| FR | France | 47.8° | 2.5° | 0.84 h | 7.91 h | 67 026 224 | 82.04 % |
| AT | Austria | 48.0° | 15.4° | −0.03 h | 7.92 h | 8 822 267 | 87.48 % |
| SK | Slovakia | 48.7° | 18.7° | −0.25 h | 7.98 h | 5 443 120 | 80.45 % |
| LU | Luxembourg | 49.6° | 6.1° | 0.59 h | 8.05 h | 602 005 | 97.06 % |
| CZ | Czech Republic | 49.9° | 15.4° | −0.02 h | 8.07 h | 10 610 055 | 80.69 % |
| BE | Belgium | 50.9° | 4.4° | 0.71 h | 8.15 h | 11 398 589 | 88.65 % |
| DE | Germany | 51.0° | 9.3° | 0.38 h | 8.17 h | 82 792 351 | 87.04 % |
| PL | Poland | 51.8° | 19.3° | −0.28 h | 8.23 h | 37 976 687 | 77.54 % |
| NL | Netherlands | 52.1° | 5.2° | 0.66 h | 8.26 h | 17 181 084 | 91.89 % |
| GB | United Kingdom | 52.1° | −1.4° | 0.09 h | 8.26 h | 66 273 576 | 90.69 % |
| IE | Ireland | 53.3° | −6.4° | 0.43 h | 8.38 h | 4 830 392 | 87.00 % |
| LT | Lithuania | 54.9° | 24.0° | 0.40 h | 8.55 h | 2 808 901 | 79.72 % |
| DK | Denmark | 55.7° | 11.6° | 0.23 h | 8.64 h | 5 781 190 | 97.32 % |
| LV | Latvia | 56.9° | 24.1° | 0.39 h | 8.80 h | 1 934 379 | 83.58 % |
| SE | Sweden | 59.2° | 15.8° | −0.05 h | 9.12 h | 10 120 242 | 89.25 % |
| EE | Estonia | 59.4° | 24.8° | 0.35 h | 9.15 h | 1 319 133 | 89.36 % |
| FI | Finland | 61.2° | 24.9° | 0.34 h | 9.48 h | 5 513 130 | 88.89 % |

Table S1 The European Union Member States at the time of the 2018 public consultation with geographical coordinates: latitude $\phi$, longitude $\lambda$, time offset —the difference between noon and midday, $Z-\lambda/\Omega$, where $Z$ is the time zone offset and $\Omega$ is Earth's rotation speed—, the winter sunrise time SRW (mean solar time), 2018 population numbers from Eurostat `demo_pjanind.tsv` table, and the internet usage from the World Bank `https://data.worldbank.org/indicator/IT.NET.USER.ZS`.

| Country | | Q2 Cancel Absolute | $C$ | | Q2 Keep Absolute | $K$ | | Net $C-K$ | | Turnout $C+K$ | | Ratio $C/K$ | |
|---|---|---|---|---|---|---|---|---|---|---|---|---|---|
| CY | Cyprus | 3557 | 0.487 % | | 3951 | 0.541 % | (3.5) | −0.054 % | | 1.029 % | | 0.900 | |
| MT | Malta | 625 | 0.161 % | | 537 | 0.138 % | | 0.023 % | | 0.299 % | | 1.164 | |
| GR | Greece | 15 829 | 0.204 % | | 20 443 | 0.263 % | | −0.059 % | | 0.467 % | | 0.774 | |
| PT | Portugal | 29 045 | 0.378 % | | 5223 | 0.068 % | | 0.310 % | | 0.446 % | | 5.561 | |
| ES | Spain | 81 961 | 0.204 % | | 5971 | 0.015 % | | 0.189 % | | 0.219 % | | 13.727 | (1.9) |
| BG | Bulgaria | 11 008 | 0.241 % | | 2123 | 0.046 % | | 0.195 % | | 0.288 % | | 5.185 | |
| IT | Italy | 15 464 | 0.034 % | | 7976 | 0.018 % | | 0.017 % | | 0.052 % | | 1.939 | |
| HR | Croatia | 19 493 | 0.631 % | | 2284 | 0.074 % | | 0.557 % | | 0.704 % | | 8.535 | |
| RO | Romania | 5827 | 0.042 % | | 1663 | 0.012 % | | 0.030 % | | 0.054 % | | 3.504 | |
| SI | Slovenia | 13 177 | 0.799 % | | 1911 | 0.116 % | | 0.683 % | | 0.915 % | | 6.895 | |
| HU | Hungary | 18 203 | 0.245 % | | 1952 | 0.026 % | | 0.218 % | | 0.271 % | | 9.325 | |
| FR | France | 328 124 | 0.597 % | | 64 497 | 0.117 % | | 0.479 % | | 0.714 % | | 5.087 | |
| AT | Austria | 200 160 | 2.594 % | (4.7) | 58 530 | 0.758 % | (5.5) | 1.835 % | (3.0) | 3.352 % | (4.5) | 3.420 | |
| SK | Slovakia | 26 435 | 0.604 % | | 6447 | 0.147 % | | 0.456 % | | 0.751 % | | 4.100 | |
| LU | Luxembourg | 8337 | 1.427 % | (1.9) | 2196 | 0.376 % | (2.1) | 1.051 % | | 1.803 % | (1.7) | 3.796 | |
| CZ | Czech Republic | 52 233 | 0.610 % | | 10 391 | 0.121 % | | 0.489 % | | 0.731 % | | 5.027 | |
| BE | Belgium | 52 267 | 0.517 % | | 10 143 | 0.100 % | | 0.417 % | | 0.618 % | | 5.153 | |
| DE | Germany | 2 633 311 | 3.654 % | (7.3) | 502 972 | 0.698 % | (4.9) | 2.956 % | (5.6) | 4.352 % | (6.3) | 5.236 | |
| PL | Poland | 121 668 | 0.413 % | | 6306 | 0.021 % | | 0.392 % | | 0.435 % | | 19.294 | (3.5) |
| NL | Netherlands | 21 851 | 0.138 % | | 5948 | 0.038 % | | 0.101 % | | 0.176 % | | 3.674 | |
| GB | United Kingdom | 9582 | 0.016 % | | 2117 | 0.004 % | | 0.012 % | | 0.019 % | | 4.526 | |
| IE | Ireland | 10 205 | 0.243 % | | 1436 | 0.034 % | | 0.209 % | | 0.277 % | | 7.107 | |
| LT | Lithuania | 8744 | 0.390 % | | 833 | 0.037 % | | 0.353 % | | 0.428 % | | 10.497 | |
| DK | Denmark | 5042 | 0.090 % | | 1196 | 0.021 % | | 0.068 % | | 0.111 % | | 4.216 | |
| LV | Latvia | 6448 | 0.399 % | | 1146 | 0.071 % | | 0.328 % | | 0.470 % | | 5.627 | |
| SE | Sweden | 42 562 | 0.471 % | | 5815 | 0.064 % | | 0.407 % | | 0.536 % | | 7.319 | |
| EE | Estonia | 10 561 | 0.896 % | | 1868 | 0.158 % | | 0.737 % | | 1.054 % | | 5.654 | |
| FI | Finland | 50 288 | 1.026 % | | 2672 | 0.055 % | | 0.972 % | | 1.081 % | | 18.820 | (3.3) |
| Median (all) | | | 0.406 % | | | 0.069 % | | 0.341 % | | 0.469 % | | 5.169 | |
| Average (w/o outliers) | | | 0.393 % | | | 0.074 % | | 0.330 % | | 0.486 % | | 4.969 | |
| Std dev (w/o outliers) | | | 0.273 % | | | 0.062 % | | 0.298 % | | 0.321 % | | 2.453 | |
| StdDev/Average | | | 69 % | | | 84 % | | 90 % | | 66 % | | 49 % | |
| $p-$val normal (all) | | | 0.02 | | | 0.01 | | 0.06 | | 0.03 | | 0.08 | |
| $p-$val normal (w/o outliers) | | | 0.60 | | | 0.48 | | 0.89 | | 0.82 | | 0.58 | |

Table S2 The results to question 2 in the 2018 public consultation conducted by the European Commission. Member States are listed in increasing values of latitude. Each answer to the question is given in absolute numbers and in shares of the target population $P \times I$. The net shares $C - K$, the turnout $C + K$ and the ratio $C/K$ are shown. Tukey's fences ($k = 1.5$) were used to identify outliers, noted in blue ink. In parenthesis the distance to the third quartile in units of the interquantile range. The bottom rows display univariate descriptive statistics: the median value for the full set; the average, standard deviation and their ratio after outlier removal; the $p-$val of the null hypothesis "sample is normally distributed" for the full set and after removal of outliers.

| **Outcome** | **Pearson-**$R^2$ | $p-$val | **Slope** [95 % **CI**] | **Unbiased SE** | **Outliers** |
|---|---|---|---|---|---|
| *Predictor: Time offset* | | | | | |
| Cancel $C$ | 0.014 | 0.570 | | 70 % | AT DE LU |
| Keep $K$ | 0.001 | 0.894 | | 86 % | AT CY DE LU |
| Net $C-K$ | 0.005 | 0.732 | | 92 % | AT DE |
| Turnout $C+K$ | 0.034 | 0.377 | | 66 % | AT DE LU |
| Rate $C/K$ | 0.001 | 0.873 | | 50 % | FI PL ES |
| *Predictor:* SRW | | | | | |
| Cancel $C$ | 0.177 | 0.036 | 0.185[0.013, 0.357] | 64 % | AT DE LU |
| Keep $K$ | 0.030 | 0.420 | | 85 % | AT CY DE LU |
| Net $C-K$ | 0.247 | 0.010 | 0.244[0.064, 0.424] | 80 % | AT DE |
| Turnout $C+K$ | 0.057 | 0.252 | | 66 % | AT DE LU |
| Rate $C/K$ | 0.239 | 0.013 | 2.248[0.517, 3.978] | 44 % | FI PL ES |

Table S3 The association between the geographical data (predictors) and the results to question 2 in the 2018 public consultation conducted by the European Commission. The null hypothesis fails to sustain for SRW and $C$, $C-K$, $C/K$ and sustains for SRW and $C+K$. The null hypothesis always fails when time offset is the predictor. The unbiased standard error (SE) is scaled by the sample average value of the either outcome. The null hypothesis of normally distributed residuals sustained in every regression at the standard level of significance. The slopes are given in basis points per hour. The positive value shows larger shares of $C$, $C-K$ and $C/K$ with higher SRW.

| Country | | Round | Sleep and other personal cares | | | | Work | | | |
|---|---|---|---|---|---|---|---|---|---|---|
| | | | offset | | midpoint | | onset | | midpoint | |
| | | | Local time | Distance to winter sunrise | Local time | Distance to winter sunrise | Local time | Distance to winter sunrise | Local time | Distance to winter sunrise |
| GR | Greece | 2 | 07:40h | −00:00h | 04:10h | −03:30h | 08:00h | +00:19h | 12:40h | +04:59h |
| ES | Spain | 2 | 08:20h | −00:12h | 04:20h | −04:12h | 08:20h | −00:12h | 13:00h | +04:27h |
| BG | Bulgaria | 1 | 07:10h | −00:39h | 03:20h | −04:29h | 08:00h | +00:10h | 13:00h | +05:10h |
| IT | Italy | 2 | 07:30h | −00:16h | 03:30h | −04:16h | 08:00h | +00:13h | 12:20h | +04:33h |
| RO | Romania | 2 | 07:20h | −00:37h | 03:10h | −04:47h | 08:00h | +00:02h | 12:10h | +04:12h |
| SI | Slovenia | 1 | 06:50h | −00:53h | 02:50h | −04:53h | 07:00h | −00:43h | 12:00h | +04:16h |
| HU | Hungary | 2 | 07:00h | −00:29h | 02:50h | −04:39h | 07:40h | +00:10h | 12:00h | +04:30h |
| FR | France | 2 | 07:50h | −00:50h | 03:40h | −05:00h | 08:00h | −00:40h | 13:20h | +04:39h |
| LU | Luxembourg | 2 | 07:40h | −00:51h | 03:40h | −04:51h | 08:00h | −00:31h | 12:50h | +04:18h |
| BE | Belgium | 2 | 07:50h | −00:54h | 03:40h | −05:04h | 08:20h | −00:24h | 12:50h | +04:05h |
| DE | Germany | 2 | 07:30h | −00:53h | 03:20h | −05:03h | 07:50h | −00:33h | 12:10h | +03:46h |
| PL | Poland | 2 | 07:20h | −00:25h | 03:00h | −04:45h | 07:20h | −00:25h | 12:20h | +04:34h |
| NL | Netherlands | 2 | 08:00h | −00:48h | 03:50h | −04:58h | 08:20h | −00:28h | 13:00h | +04:11h |
| GB | United Kingdom | 2 | 07:50h | −00:26h | 03:30h | −04:46h | 08:30h | +00:13h | 13:00h | +04:43h |
| LT | Lithuania | 1 | 07:00h | −01:49h | 02:50h | −05:59h | 07:40h | −01:09h | 12:50h | +04:00h |
| LV | Latvia | 1 | 07:20h | −01:42h | 03:10h | −05:52h | 08:00h | −01:02h | 13:20h | +04:17h |
| SE | Sweden | 1 | 07:30h | −01:17h | 03:20h | −05:27h | 07:50h | −00:57h | 12:40h | +03:52h |
| EE | Estonia | 2 | 07:50h | −01:28h | 03:40h | −05:38h | 08:00h | −01:18h | 12:50h | +03:31h |
| FI | Finland | 2 | 07:40h | −01:59h | 03:30h | −06:09h | 08:00h | −01:39h | 12:30h | +02:50h |

Table S4 The time marks associated with the sleep and other personal cares and the work cycles as per HETUS. Countries are listed in increasing values of latitude. The third left-most column list the round of HETUS: round 1 was taken around 2000; round 2 around 2010. Round 3 is currently in progress. Figure S3 shows the methodology to asses midpoints and offset/onset time marks.

| **Predictor** | **Pearson-$R^2$** | $p-$val | **Slope**[95 % **CI**] | **Unbiased SE** |
|---|---|---|---|---|
| | | *Sleep and other personal cares offset* | | |
| Distance to solar noon | 0.007 | 0.751 | | 80 % |
| Distance to SRW | 0.447 | 0.003 | −0.34[−0.55, −0.13] | 59 % |
| | | *Work onset* | | |
| Distance to solar noon | 0.161 | 0.111 | | 73 % |
| Distance to SRW | 0.677 | $5 \times 10^{-5}$ | −0.40[−0.56, −0.25] | 45 % |
| | | *Sleep and other personal cares midpoint* | | |
| Distance to solar noon | 0.031 | 0.500 | | 79 % |
| Distance to SRW | 0.376 | 0.009 | −0.27[−0.46, −0.08] | 63 % |
| | | *Work midpoint* | | |
| Distance to solar noon | 0.001 | 0.918 | | 80 % |
| Distance to SRW | 0.474 | 0.002 | −0.38[−0.60, −0.16] | 58 % |
| Geographical predictors | | | | |
| Time offset | 0.000 | 0.960 | | 80 % |
| SRW | 0.384 | 0.008 | 0.29[0.09, 0.50] | 63 % |

Table S5 The association between time marks of human activity —measured through two distinct metrics— (predictor, see Table S4) and the shares of target population willing to cancel the time regulations as per the 2018 public consultation (outcome, see Table S2) in $N = 17$ Member States that reported Hetus statistics. Blue ink annotates $p-$values below the standard level of significance ($\alpha = 0.05$), which occurs only when predictors are measured relative to the winter sunrise. The negative slope shows that shares against the current regulations increases with earlier starting points relative to winter sunrise time. Slopes are given in basis points per hour. The unbiased standard error of the fit is scaled by sample average value of the fitted shares $C$ (0.390 %) and the result expressed as a percentage. The null hypothesis of normally distributed residuals sustains in every regression at the standard level of significance. For comparison the association with geographical predictions in this subset is also shown.

#### 2. ADDITIONAL FIGURES

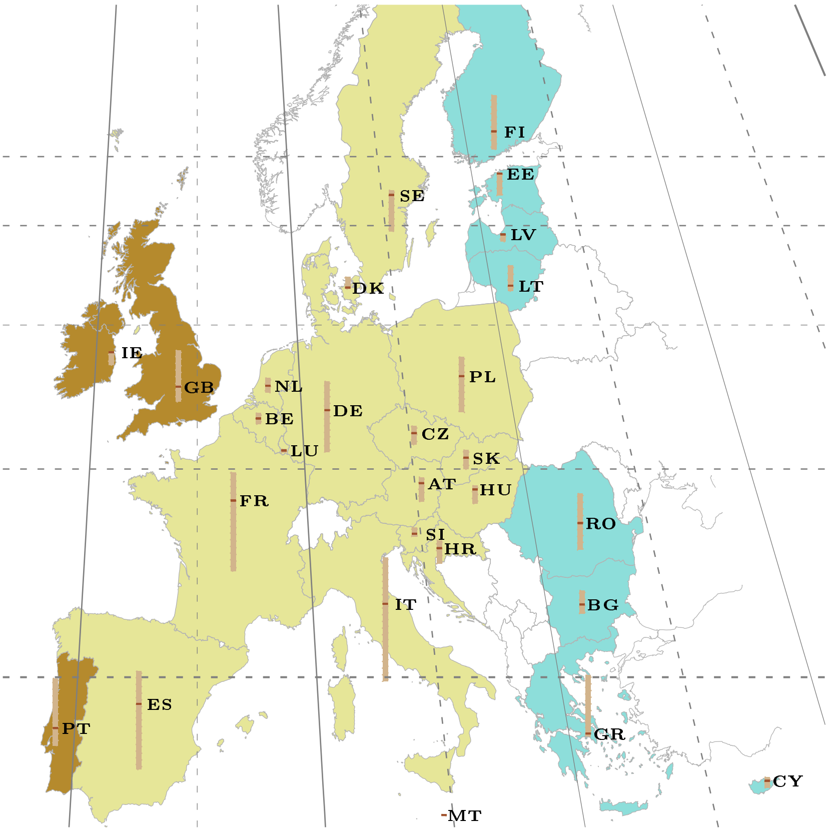

Figure S1 An Albers projection of Europe with the countries participating in the 2018 public consultation on summertime arrangements (in shade and labeled). Dashed meridians annotate time meridians. Solid meridians annotate the bounduaries of the physical time zones. Circles of latitudes annotate straight values of SRW, starting at SRW = 7.5 h (southern most) and ending at 9.5 h (northern most) in steps of 0.5 h. Color shades highlight time zones; from west to east, Western European Time ($Z = 0$), Central European Time ($Z = 1$ h) and Eastern European Time ($Z = 2$ h). Vertical bar annotates the quartile range of population weighted latitude at the median population weighted latitude. The black horizontal bar locates the pupulation median latitude and longitude for the country, listed in Table S1. The map was made with Natural Earth, a free vector and raster map data available at `https://www.naturalearthdata.com/`.

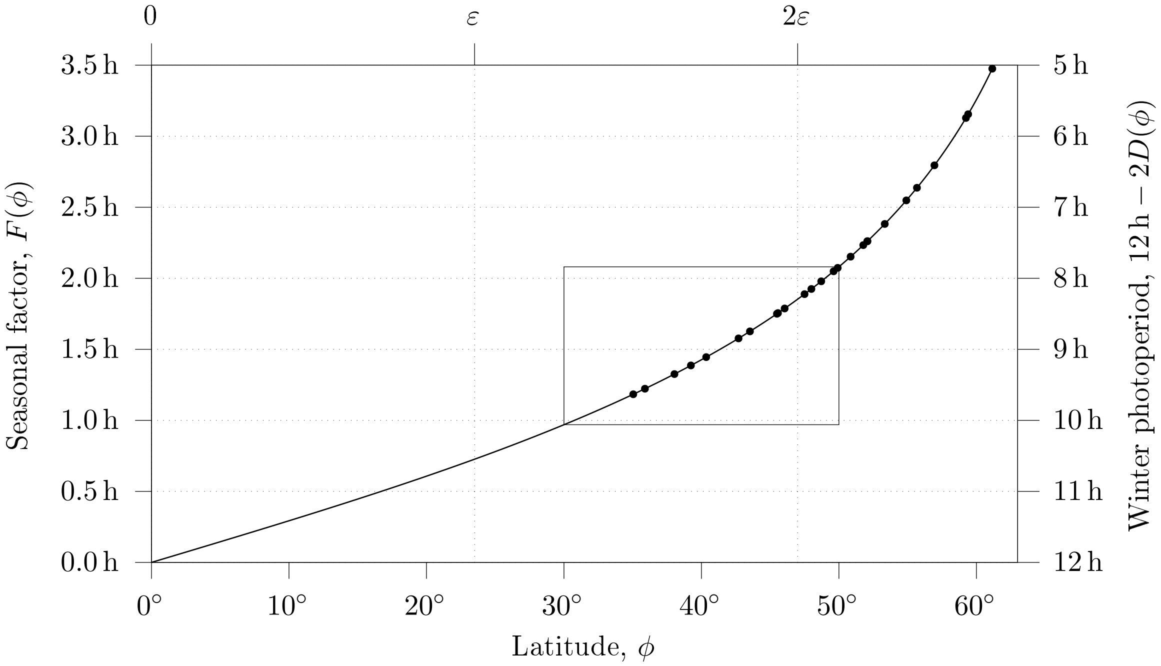

Figure S2 The evolution of the seasonal factor $F$ as a function of latitude $\phi$. The seasonal factor is defined in Eq. (**??**) so that it equals to zero at the Equator. The seasonal factor gives the advance or delay of the sunrise and sunset times at the solstices relative to the Equatorial standard. The right axis shows the corresponding winter photoperiod (and the summer scotoperiod) $12\,\mathrm{h} - 2F(\phi)$. Data points annotate the Member States. The seasonal factor was modeled for a point-sized Sun and a refractionless atmosphere. The box expands from $30^\circ$ to $50^\circ$ and yields $F(\phi)$ from $1\,\mathrm{h}$ to $2\,\mathrm{h}$. It is an educated guess for the utility range of seasonal regulations of time.

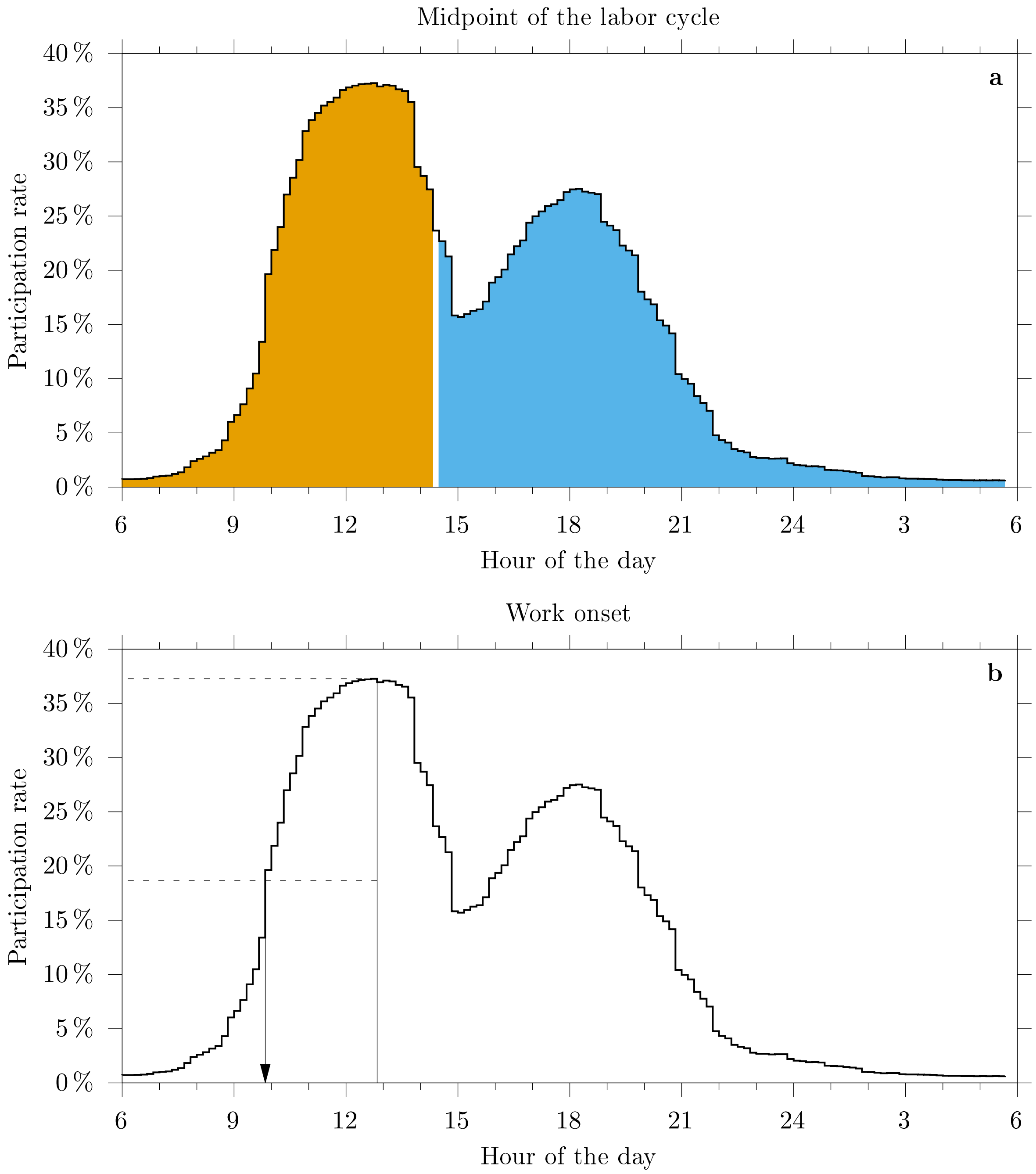

Figure S3 Determination of the midpoint and work onset. On the left panel the daily rhythm of labor as per HETUS. The area enclosed by the daily rhythm scales with the daily average total work. The shaded areas break even the total area. The hour of the day when this happens is the *midpoint* of the labor cycle. On the right panel, the maximum participation rate is halved. Work onset is determined as the hour of the day when the participation rate first exceeds the maximum daily participation rate. Similar analysis allow to assess the midpoint of the sleep/wake cycle and the sleep offset.

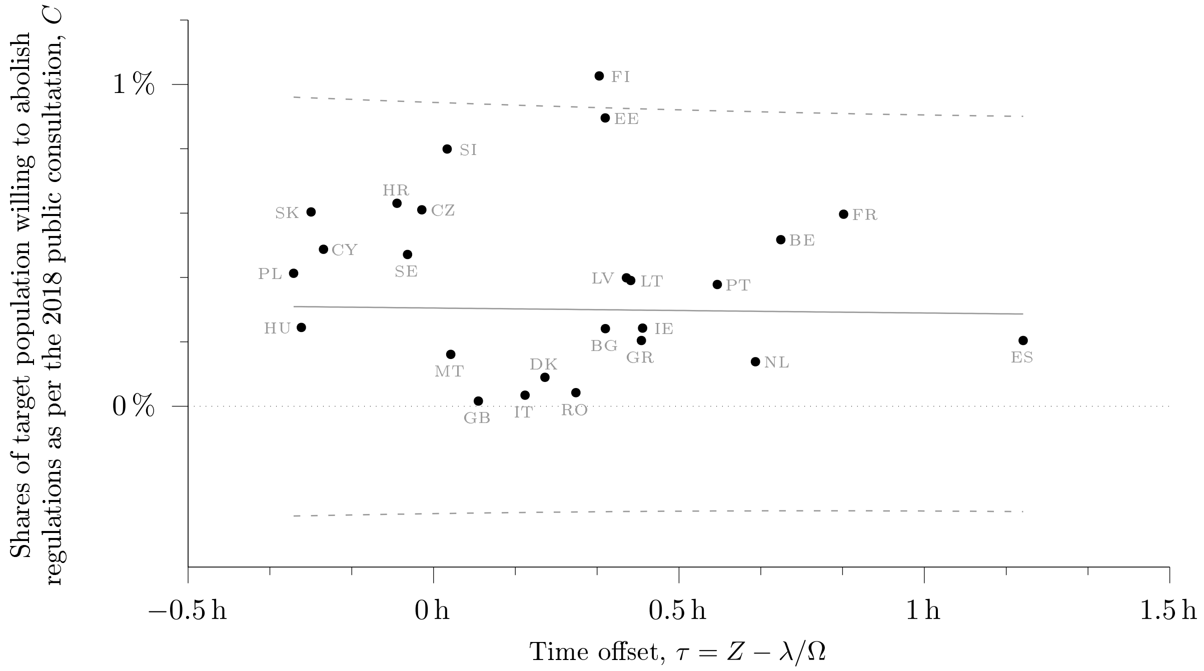

Figure S4 The association between the time offset —difference between noon and midday— and the shares of target population willing to cancel the regulations as per the 2018 public consultation $C$. Germany (3.18 %), Austria (2.27 %) and Luxembourg (1.38 %) are not shown in the picture. The solid line shows the point estimates of the prediction; the dashed lines bound the 95 % prediction interval. The null hypothesis "shares $C$ do not depend on time offset" sustains ($p = 0.696$ at the standard level of significance ($\alpha = 0.05$), see Table S3 in the Supplemental Information. Labels show iso-3166 alpha-2 codes, see table S1.

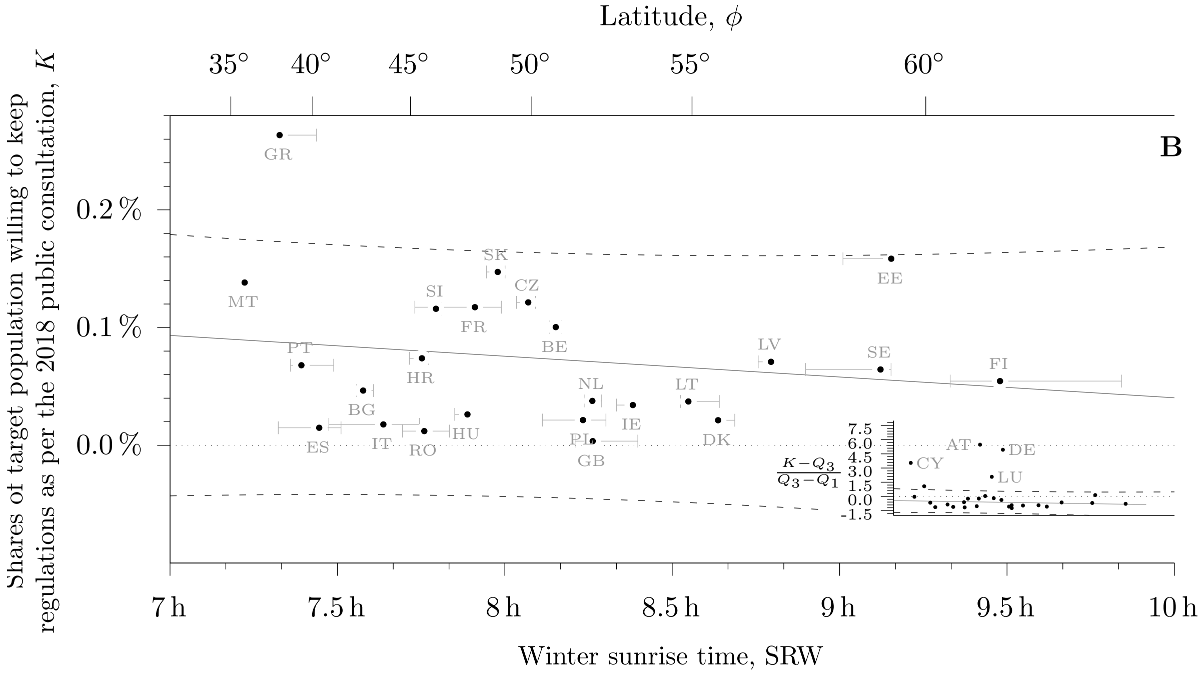

Figure S5 The same as Figure 1B (main file) but in a magnified vertical scale to visualize the distribution of shares against the regulations.

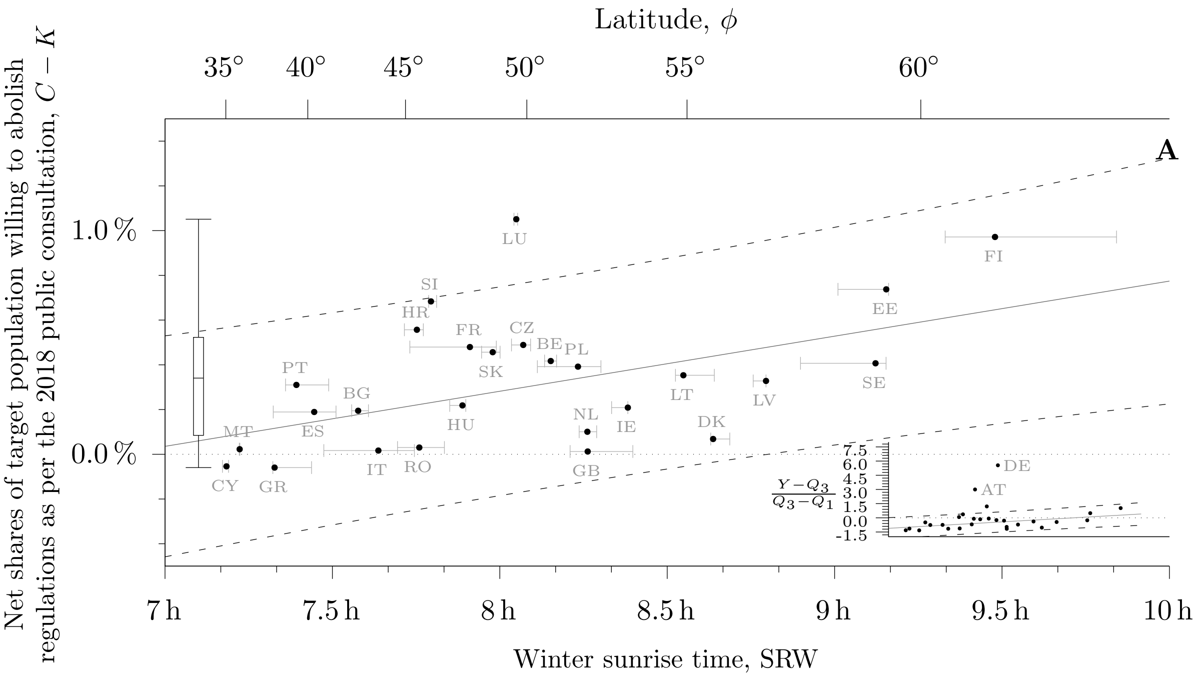

Figure S6 The association between the winter sunrise time SRW and the net shares of population willing to abolish DST $C-K$ as per the 2018 public consultation $C$. The inset shows the distribution of shares scaled by the interquartile range to identify and quantify the outliers Austria, Germany, not shown in the main picture and excluded from the regression. The low quartile was $Q_1 = 0.085\,\%$ and the high quartile, $Q_3 = 0.522\,\%$ (see the boxplot on the left side of the plot). The solid line shows the point estimates of the prediction; the dashed lines bound the 95 % prediction interval. The regression is statistically significant ($p = 0.010$) at the standard level of significance ($\alpha = 0.05$). Notice that the span of the predictor is larger than one hour: the standard unit of social time and the size of the shift brought by DST to clocks. Horizontal bars highlight population weighted low and high quartiles (France and Spain show the span of the European areas only). Labels (iso-3166 alpha-2) in increasing values of latitude: CY Cyprus; MT Malta; GR Greece; PT Portugal; ES Spain; BG Bulgaria; IT Italy; HR Croatia; RO Romania; SI Slovenia; HU Hungary; FR France; AT Austria; SK Slovakia; LU Luxembourg; CZ Czech Republic; BE Belgium; DE Germany; PL Poland; GB United Kingdom; NL Netherlands; IE Ireland; LT Lithuania; DK Denmark; LV Latvia; SE Sweden; EE Estonia; FI Finland.

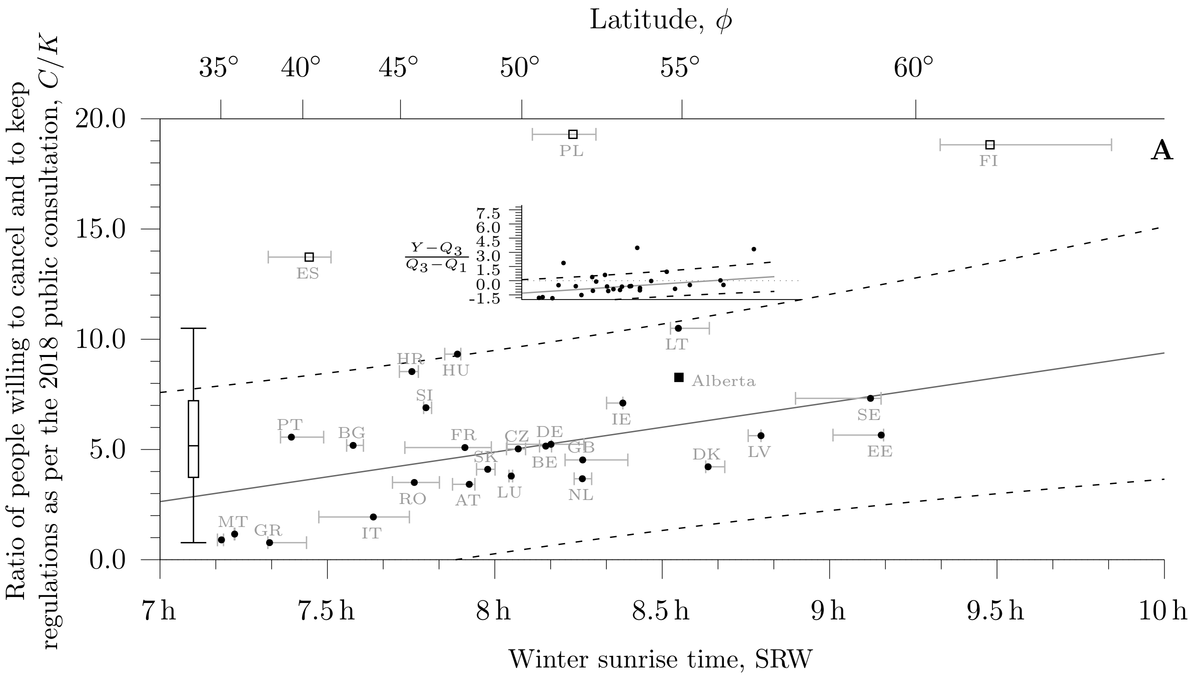

Figure S7 The association between the winter sunrise time SRW and the ratio of shares of population willing to cancel and keep DST $C/K$ as per the 2018 public consultation $C$. The results from the 2019 public consultation in Alberta (Canada) —permanent DST vs current regulations— are also shown. The inset shows the distribution of shares scaled by the interquartile range to identify and quantify the outliers Austria, Germany, not shown in the main picture and excluded from the regression. The low quartile was $Q_1 = 3.735$ and the high quartile, $Q_3 = 7.213$ (see the boxplot on the left side of the plot). The solid line shows the point estimates of the prediction; the dashed lines bound the 95 % prediction interval. The regression is statistically significant ($p = 0.013$) at the standard level of significance ($\alpha = 0.05$). Notice that the span of the predictor is larger than one hour: the standard unit of social time and the size of the shift brought by DST to clocks. Horizontal bars highlight population weighted low and high quartiles (France and Spain show the span of the European areas only). Labels (iso-3166 alpha-2) in increasing values of latitude: CY Cyprus; MT Malta; GR Greece; PT Portugal; ES Spain; BG Bulgaria; IT Italy; HR Croatia; RO Romania; SI Slovenia; HU Hungary; FR France; AT Austria; SK Slovakia; LU Luxembourg; CZ Czech Republic; BE Belgium; DE Germany; PL Poland; GB United Kingdom; NL Netherlands; IE Ireland; LT Lithuania; DK Denmark; LV Latvia; SE Sweden; EE Estonia; FI Finland.

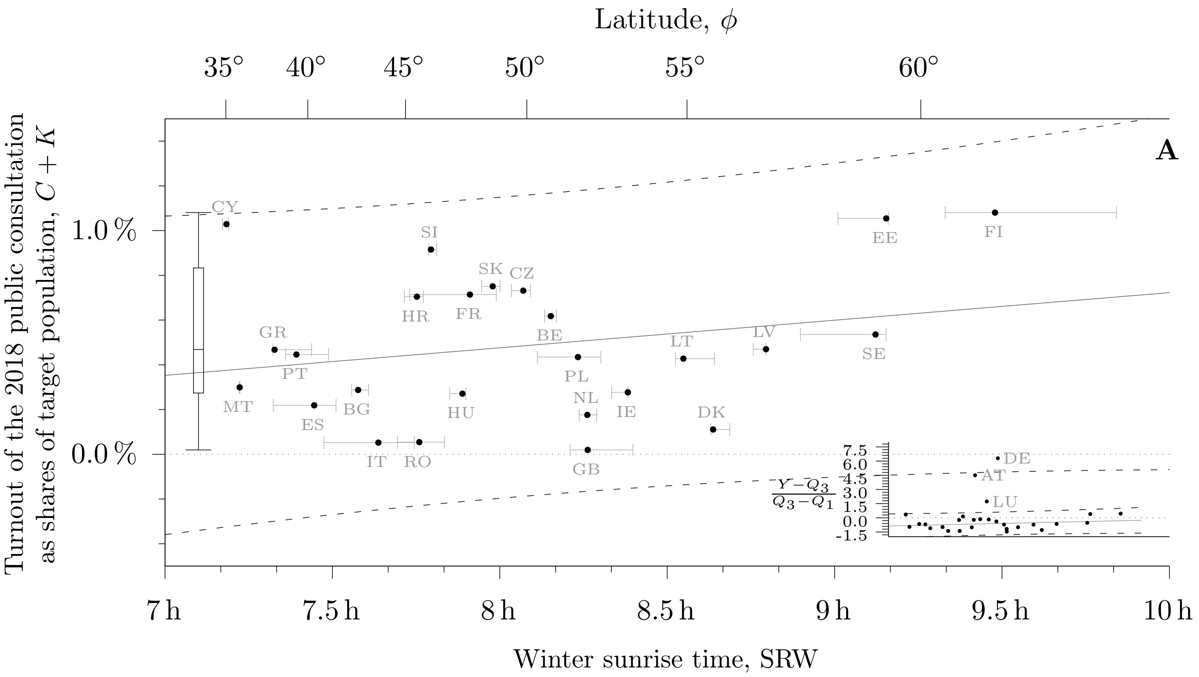

Figure S8 The association between the winter sunrise time SRW and the turnout $C + K$ to question 2 from the 2018 public consultation. The inset shows the distribution of shares scaled by the interquartile range to identify and quantify the outliers Austria, Germany, not shown in the main picture and excluded from the regression. The low quartile was $Q_1 = 0.274\,\%$ and the high quartile, $Q_3 = 0.833\,\%$ (see the boxplot on the left side of the plot). The solid line shows the point estimates of the prediction; the dashed lines bound the 95 % prediction interval. The regression is not statistically significant ($p = 0.252$) at the standard level of significance ($\alpha = 0.05$). Notice that the span of the predictor is larger than one hour: the standard unit of social time and the size of the shift brought by DST to clocks. Horizontal bars highlight population weighted low and high quartiles (France and Spain show the span of the European areas only). Labels (iso-3166 alpha-2) in increasing values of latitude: CY Cyprus; MT Malta; GR Greece; PT Portugal; ES Spain; BG Bulgaria; IT Italy; HR Croatia; RO Romania; SI Slovenia; HU Hungary; FR France; AT Austria; SK Slovakia; LU Luxembourg; CZ Czech Republic; BE Belgium; DE Germany; PL Poland; GB United Kingdom; NL Netherlands; IE Ireland; LT Lithuania; DK Denmark; LV Latvia; SE Sweden; EE Estonia; FI Finland.

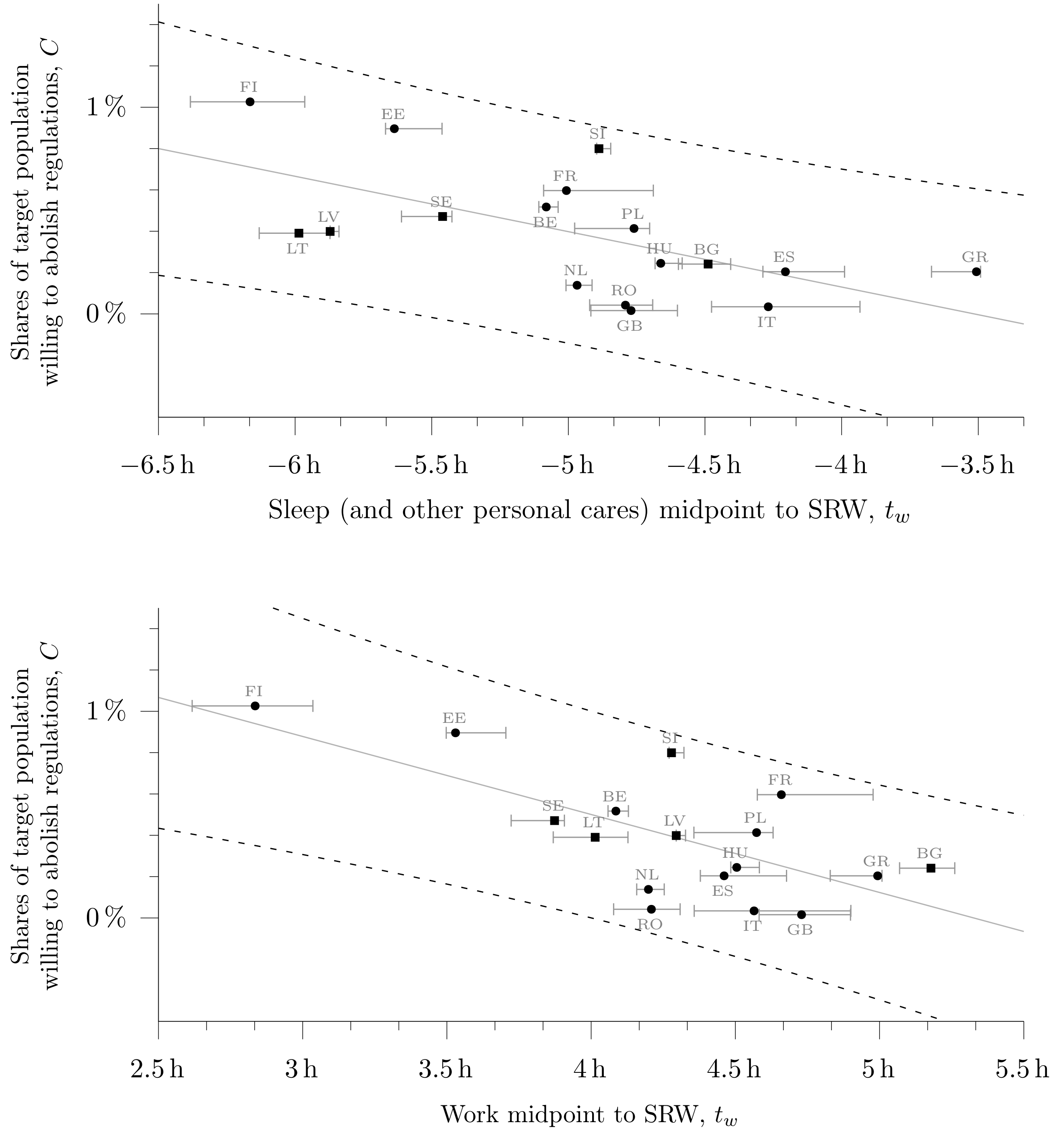

Figure S9 Scatter plots of the shares of target population willing to cancel DST as per the 2018 European Commission public consultation (outcome) vs two predictors related to human activity as per HETUS: midpoint of sleep to SRW (top) and midpoint of wokr to SRW (bottom). Circles refer to round 2 (year 2010) of HETUS, and squares to round 1 (year 2000). The null hypothesis “the predictor does not explain the outcome” does not sustain at the standard level of significance for either test. For midpoint of sleep $R^2 = 0.376, p = 0.009, N = 17$; for the midpoint of work $R^2 = 0.474, p = 0.002, N = 17$. Solid lines show the results of the linear regression; dashed lines bound the 95 % prediction interval of the regression. Horizontal bars highlight population weighted low and high quartiles (France and Spain show the span of the European areas only). The null hypotheses sustained if the descriptor is set to distance to noon.
